# Assessing the Impact of Human Mobility to Predict Regional Excess Death in Ecuador

**DOI:** 10.1101/2021.05.19.21257056

**Authors:** Leticia Cuéllar, Irene Torres, Ethan Romero-Severson, Riya Mahesh, Nathaniel Ortega, Sara Pungitore, Ruian Ke, Nicolas Hengartner

## Abstract

COVID-19 outbreaks have had high mortality in low- and medium-income countries such as Ecuador. Human mobility is an important factor influencing the spread of diseases possibly leading to a high burden of disease at the country level. Drastic control measures, such as complete lockdown are effective epidemic controls, yet in practice, one hopes that a partial shutdown would suffice. It is an open problem to determine how much mobility can be allowed while controlling an outbreak. In this paper, we use statistical models to relate human mobility to the excess death in Ecuador while controlling for demographic factors. The mobility index provided by GRANDATA, based on mobile phone users, represents the change of number of out-of-home events with respect to a benchmark date (March 2^nd^, the first date the data is available). The study confirms the global trend that more men are dying than expected compared to women, and that people under 30 show less deaths than expected. Specifically, individuals in the age groups younger than 20, we found have their death rate reduced during the pandemic between 22% and 27% of the expected deaths in the absence of COVID-19. The weekly median mobility time series shows a sharp decrease in human mobility immediately after a national lockdown was declared on March 17, 2020 and a progressive increase towards the pre-lockdown level within two months. Relating median mobility to excess death shows a lag in its effect: first, a decrease in mobility in the previous two to three weeks decreases the excess death and more novel, we found that an increase of mobility variability four weeks prior, increases the number of excess deaths.

## 1. Introduction

The coronavirus disease (COVID-19) pandemic has a high morbidity and mortality. Ecuador, like many Latin American countries, has been hit hard by the COVID-19 pandemic, with over 242 thousand reported cases and 14,500 deaths by the end of January 2021. The first confirmed case of COVID-19, reported on February 29, 2020, was a woman in her 70’s who returned from Spain two weeks prior [8]. On March, 13, with 23 confirmed cases, that same woman became the first COVID-19 confirmed fatality [19]. By April, Ecuador emerged as an “epicenter” of the pandemic in Latin America, with reports of uncollected dead bodies remaining for days in the streets [15]. To control the outbreak, schools in Ecuador were closed on March 13th, and on March 17th, Ecuador implemented a national lock down. Both of these measures decrease the spread of the disease by reducing contacts between infected and susceptible individuals. It is possible to measure the compliance to these orders by tracking human mobility derived from cell phone data. In this paper, we empirically address the hypothesis that a reduction in mobility predicts a decrease in the future number of COVID-19 cases and deaths.

Quantifying the severity of the outbreak in Ecuador is challenging. Due to limited testing, the reported daily counts of COVID-19 incidence and death underestimate the true magnitude of the outbreak. Excess death, which compares total number of observed deaths to the expected number of deaths, is commonly used to assess official undercounted burden of an infectious disease. From a health care and societal perspective, quantifying the excess death associated to an epidemic is informative [1], [47], [48], and is the most reliable measure of current COVID-19 data available [24]. Analysis of the increase in all-cause mortality can complement the more traditional analysis of the time series of disease incidence and disease-related deaths.

Our previous analysis of excess deaths revealed the true impact of COVID-19 on mortality in Ecuador [12]. Key findings include 70% more deaths than expected during January 1^st^ to September 23^rd^, 2020, which is 3 times the level of excess deaths found in high income counties like the United States [5], [35], [39], [47], England Wales [1], and Italy [24], [38]. Strikingly, the Provinces of Guayas and Santa Elena, the worst affected by the pandemic, have more than 200% more deaths than expected, with the highest peaks in late March and early April reaching between a 12 to 15-fold more deaths than expected. As for demographics, we found similar patterns to those observed in other countries, for example we found that that men had a death rate 183% of the expected value, while the rate in women was 153% of expected deaths; and even though we find that excess death increased with age, the group mostly affected is the [60, 70) age group with a death rate 233% of the expected value, while the age group with people older than 80, that had a death rate 160% of the expected level. Finally, we found that the indigenous ethnic group was disproportionally affected with a death rate 220% more than expected compared to 136% for the mestizo’s group, the most prevalent ethnic group in Ecuador.

In this paper, we investigate the temporal dynamic of excess deaths within each province in Ecuador, and investigate how that dynamic is linked to human mobility characteristics derived from mobile phones data provided by GRANDATA and the United Nations Development Program. GRANDATA collects geolocation events of smartphone owners using a MADID (Mobile Advertising ID) “hash”. These events track the users’ mobility patterns for a sizeable sample of the population of twelve Latin American and Caribbean countries, including Ecuador. Aggregation of that data, to preserve the anonymity of the users, has emerged as a useful tool to measure human mobility and its relationship to the spread of diseases [7], [19], [43], including malaria [36], [40], cholera [31], measles [44], dengue [13], [42], and Ebola [30], [41].

To relate the dynamic of excess death to mobility, we estimate the time series of weekly all cause excess death in each province stratified by age group and sex, and calculate at the province level human mobility characteristics derived from mobile phones data provided by GRANDATA and the United Nations Development Program.

In this paper, we use the mobility data to quantify compliance with mandatory lockdown orders in Ecuador, and mobility patterns after the strict lockdown is lifted. Cellular phone mobility data are readily available, and can be aggregated at relevant geographical resolution. In our case, we have mobility data for each Ecuadorian canton. While this type of data can be collected systematically, their aggregation may limit their usefulness as a predictor of disease spread. For example, the data lacks demographic characteristics of the users. Since age and sex are important predictors for excess death [11], having the mobility data disaggregated similarly would be useful to understand how mobility [26] impacts excess death. Further, the GRANDATA excludes people with limited activity, essentially excluding proportion of individuals staying home, including those telecommuting. Nevertheless, the data from GRANDATA is a direct measure of the relationship between government restrictions, mobility, and invaluable to quantify compliance to mobility restrictions imposed to mitigate the spread of COVID.

Our analysis of the time series of the weekly Excess Death Factor (EDF), the ratio of observed death over expected death, reveals a wave of COVID that originated in Guayas and spread through the rest of Ecuador over a period of six months, from March 2020 to August 2020. This suggests that Ecuador suffered a single outbreak with a complex spatiotemporal pattern rather than two distinct waves. But the nature of the outbreak changed over time, possibly in response to the strict lockdown. The highest and earliest peak of over 10-fold EDF (death rate greater than 10 times the normal level) occurred in the provinces of Guayas and Santa Elena during the national lockdown. The peak of EDF in all other provinces occurred after, and showed at most a 4-fold increase. A statistical analysis also reveals that the relationship between mobility and EDF showed two archetypes, one pattern for the provinces whose peak EDF occurred during the strict lockdown, and another pattern for the provinces who reached the peak of their EDF after the conclusion of the strict lockdown.

From a methodology perspective we demonstrate that both the median mobility and the variability of mobility as measured by the Interquartile Range (IQR) are statistically significant predictors for the EDF. This suggest that future analysis of mobility data should consider using finer-scale mobility data with additional demographic covariates to increase the predictive power of mobility data.

## 2. Data

### 2.1 Background about Ecuador

Ecuador has an estimated population of 17.5 million; it is located by the Pacific Ocean, at the northwest of South America, and borders with Colombia to the north and Peru to the south and east. It is divided into 24 provinces, that are each further divided into cantons. In the World Bank classification, it is an upper middle-income country, with a Gini coefficient of 45.4 (where 100 is maximum inequality). There are currently an estimated 415,000 Venezuelan forced migrants in the country [9].

Ecuador’s health system is fragmented into a network of public and private providers paid by two institutions that give coverage in a segmented mechanism: public health insurance for 40% of the (employed or farmer) population, and the Ministry of Public Health services for the uninsured. At least 39% of total health spending in Ecuador is out-of-pocket [27] with more than half of this amount destined for medication. Life expectancy at birth for women is 79.3 and for men, 73.9 years [28].

Ecuador officially registered a relatively small number of COVID-19 deaths; yet news articles from March and April 2020 report a large number of deaths. Estimates of excess deaths in [10] are consistent with Ecuador having a high death toll.

### 2.2 Mobility Data

We obtained daily mobility change scores, aggregated at the canton level from GRANDATA and the United Nations Development Program for the period starting on March 1st, 2020 and ending on November 1st, 2020. Since our all-cause mortality data for 2020 ranges from January 1^st^ to September 23^rd^, the study period in this paper is March 1^st^ to September 23^rd^, 2020. GRANDATA collected geolocations of the mobile phone users to track their mobility patterns, using the mode location (the most frequent location) as their place of residence, and recorded the number of “out-of-home” events. These “out-of-home events” are aggregated daily at various regions levels, and they report the percentage difference of these “out-of-home” events for the region with respect to the baseline date, March 2nd, 2020. GRANDATA claims that the data comes from a sizable sample of the population but we do not know the exact size of the sample and they dropped from the study users with limited activity or data, those with less than 10 events for the day or whose events were all recorded only within an 8-hour window, leaving at least 16 hours unaccounted for.

GRANDATA provided mobility indexes for 191 cantons (out of the 218) with no data for 27 cantons. We excluded the small Ecuadorian “Not Delimited Areas” located along various provinces’ borders.

We summarized the mobility score for each province by computing the population weighted median, population weighted inter-quartile range, and a population weighted measure of skewness. Figure 2 displays the time series of the median mobility for each province, highlighting in color the statistics of the provinces Guayas, Manabí, Pichincha, and Santo Domingo. The box indicates the period of strict lockdown. A relative change of −0.5 indicates that there are half as many “out-of-home” travels, a value of 0 corresponds to normal out of home travel, and a positive value of 0.5 correspond to a 50% increase in the number of out-of-home trips.

**Figure 1.**
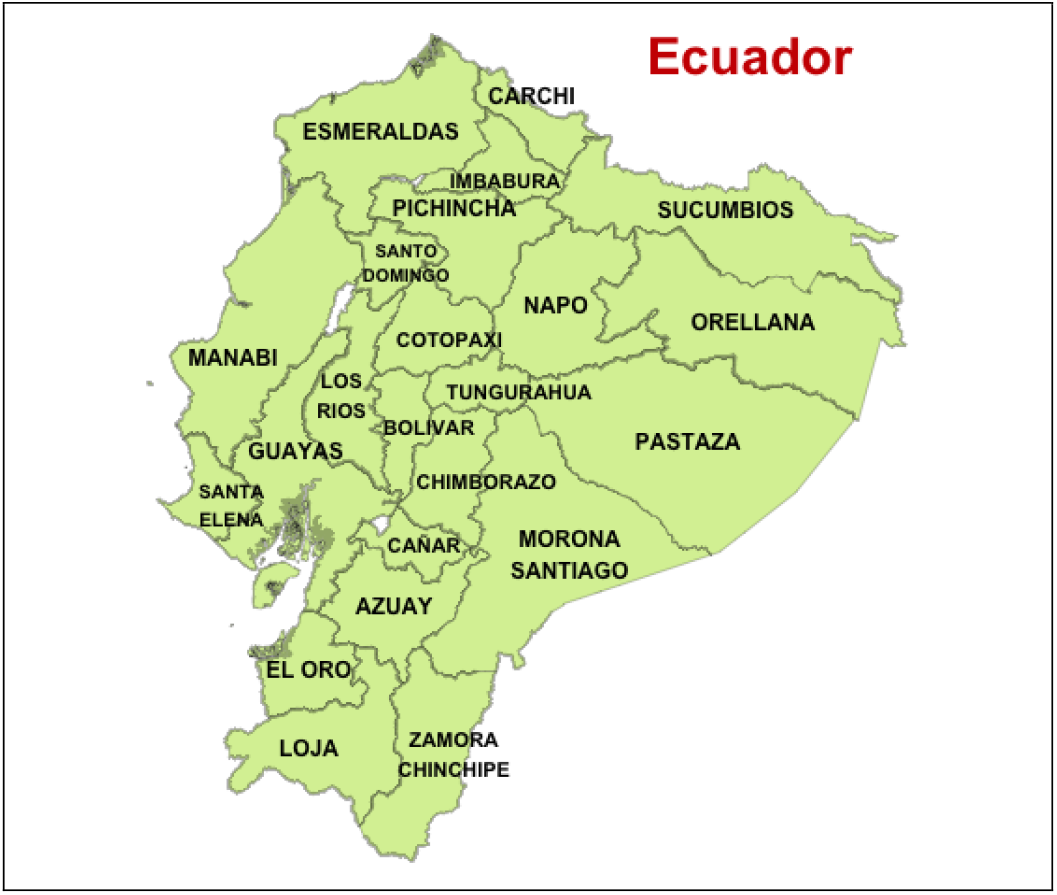
Map of Ecuador with provinces

**Figure 2.**
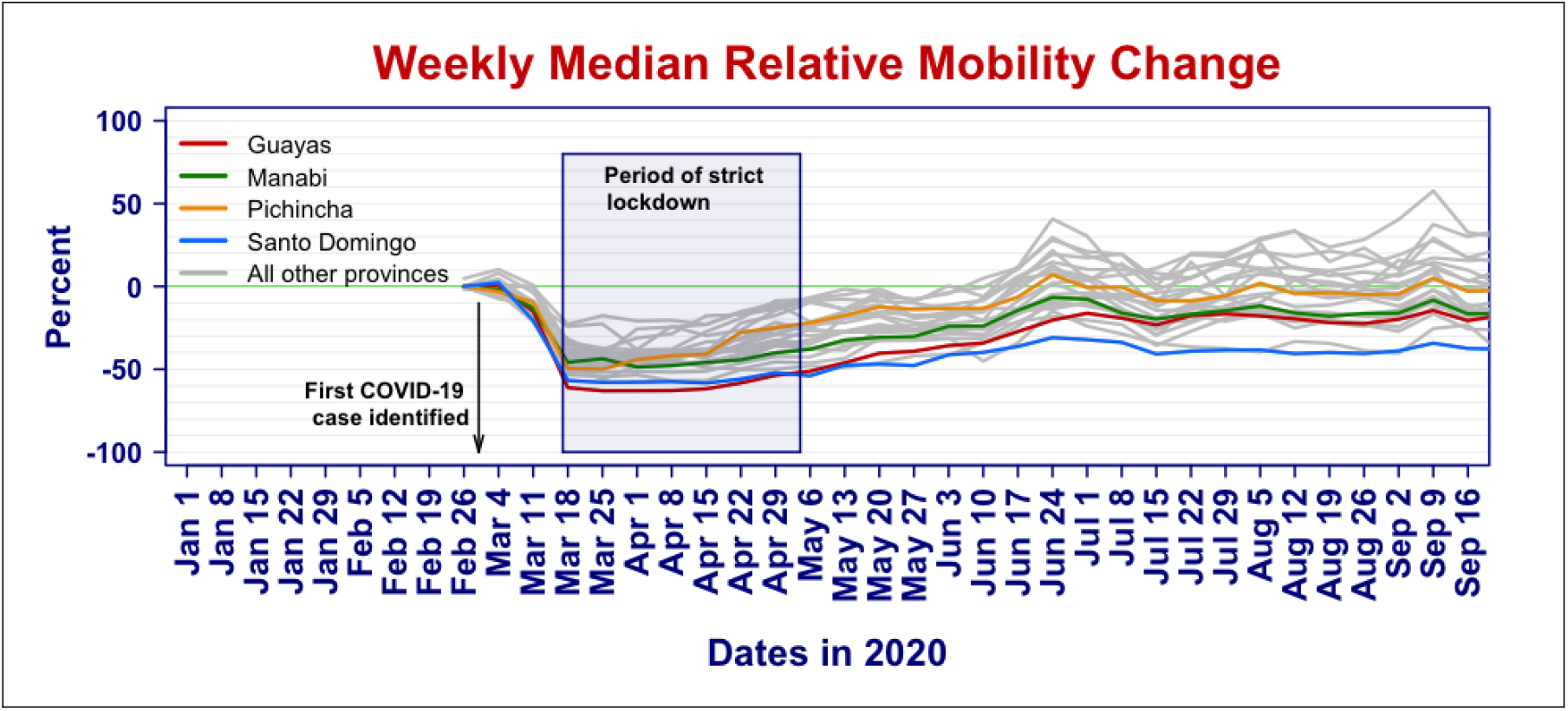
Time series of relative median mobility change for each province

We summarize the variability of the mobility scores by the Inter-Quartile Range (IQR). The IQR measures the spread of a distribution by taking the difference between the 75% quantile and the 25% quantile. For a Gaussian distribution, the IQR is 1.35 times the standard deviation. Figure 3 displays the time series of IQR in each province in gray, with the values for the provinces of Guayas, Manabí, Pichincha, and Santo Domingo highlighted in color. The box indicates the period of strict lockdown.

**Figure 3.**
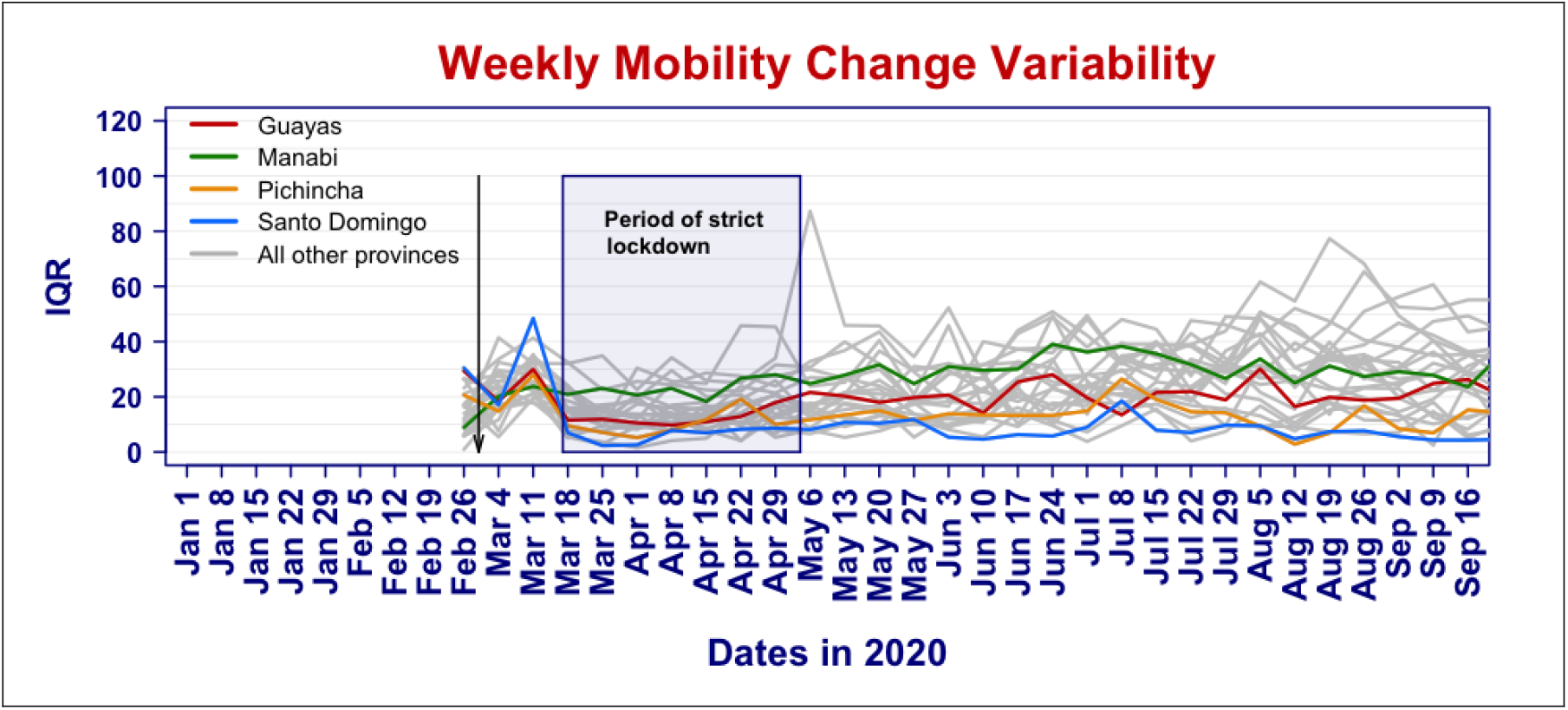
Time series of mobility change variability (IQR) for all Ecuadorian provinces

### 2.3 Death Data

Historical death records from 2015-2019 for all-cause mortality were obtained from the Ecuadorian National Institute of Statistics and Census. For each individual, the records include date of death, age, sex, and ethnicity of the diseased, place of death registration, residence, and the International Classification of Disease (ICD) code for the cause of death.

The Ecuadorian Ministry of Government provided death records from all causes that occurred from January 1st, 2020 to September 26th, 2020. In addition to the date of death, records report sex, age, registration, and residence location by parish, canton, and province, but *without the cause of death*.

For our analysis, we computed the number of death per week, sex, age, and province. We binned age into nine 10-year age groups ([0,9], [10, 19], …, [70, 80) and [80 and older]). For all years (2015-2020), week 1 was set from January 1 to January 7, week 2 from January 8 to January 14, and so on. In all cases, week 53 contains less than 7 days. We calculated counts for all 24 Ecuadorian provinces but ignored the three smaller “Not Delimited Areas”.

We use the 2020 population estimates from the INEC (the Ecuadorian National Institute of Statistics and Census).

Individual records of COVID-19 incidence and testing from Ecuador were obtained from the Ecuadorian Ministry of Public Health [6]. Death records were obtained from the Ministry of Interior. All records were aggregated at the weekly level and binned by sex, age group, and province. Ethics committee approval for the use of health patient information and death records was obtained from the Ethical Committee for Expedited Review of COVID-19 Research of the Ecuadorian Ministry of Health. The study has been approved by the Human Subjects Research Review Board, Los Alamos National Laboratory.

## Methods

### 3.1 Estimation of Excess death

We fitted a Poisson log-linear model to the aggregated past death counts using the *glm* function in R 4.0.3 and the diseased data from 2015 to 2019. The predictor variables included sex, age group and their interactions, province and week of the year as a factor. With this fitted model, we predict the time series of weekly expected number of deaths from all causes in each province, divided by age and sex. The fitting of this model was described in [12].

The excess deaths are the difference between the observed number of deaths in 2020 and the model predicted expected number of deaths; that is, they are the number of deaths above what would have been expected if 2020 were a typical year. The Excess Death Factor (EDF) is the ratio of the observed over the expected number of deaths. The EDF normalizes each province to itself making it much more interpretable than simply the number of excess deaths, which facilitates comparisons between provinces. An EDF of 2 means that there are twice as many deaths during the pandemic than in a normal year. The presumption is that these deaths are attributed to COVID-19, although there is uncertainty in that attribution. Indeed, it is possible that mortality from all causes is higher because the healthcare system is overwhelmed, or lower because the strict lockdown likely prevented some deaths (e.g. by reducing the number of vehicle accidents). However, the EDF gives a much clearer image of COVID-19 associated mortality than the confirmed COVID deaths.

### 3.2 Statistical analysis

Given the shifting delay of the outbreak in each province, we chose to model each province separately. On average, a death from COVID-19 is estimated to occur two to eight weeks after infection [45]. To explore the effect of this delay, we computed the cross-correlation between the time series of the EDF and the mobility statistics. That is, we calculated the correlation between the EDF in a given week, and the mobility statistic in prior weeks. While a correlation does not imply causation, the cross-correlation shows how changes in mobility in the past impact today’s EDF.

We used Poisson log-linear regression to explain the EDFs by mobility, while controlling for age and sex. To do so, we model the observed weekly death Y in each province as Poisson distributed variable with mean

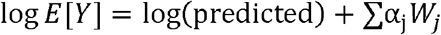

where W_1_,…, W_k_ are the explanatory variables in the model, and log(predicted) is the logarithm of the predicted baseline for the number of deaths based on the historical data. The latter is used as an offset in the regression. To account for the delay between mobility change and death, our model included lagged mobility statistics, that is, we only included mobility statistics from the 2, 3, and 4 previous weeks. Further lags were not considered for the regression analysis given the limited number of weeks for the mobility data.

We applied model selection using stepwise regression using the Akaike information criterion (AIC) criterion on the mobility statistics to identify which lags and other summary statistics were important. We note that the AIC is permissive in that it may retain a few variables that are only weakly associated with the response.

Finally, we compute a measure of variability explained by the mobility statistics by reporting the ratio of the difference of the deviance of the fitted model with and without the mobility statistics, divided by the difference of the full model [11] and the model with only the demographic variables. This quantity is similar in spirit to the R ^2^ statistic in standard linear regression.

## 4. Results

### 4.1 Geographical distribution of excess death

Figure 4 visualizes the geographical spread of the EDF for aggregated by sex and all age groups, for six selected weeks: the week of March 18, when the lock started, the week of April 1^st^, in the middle of the lockdown, the weeks of June 3^rd^, July 15 and August 16, after the lockdown, and the week of September 16, the last week we had data. A plot for all the weeks is provided in the appendix. The plot shows how the hotspot of the epidemic is moving through Ecuador, with different provinces becoming the weekly hotspot at different times.

**Figure 4.**
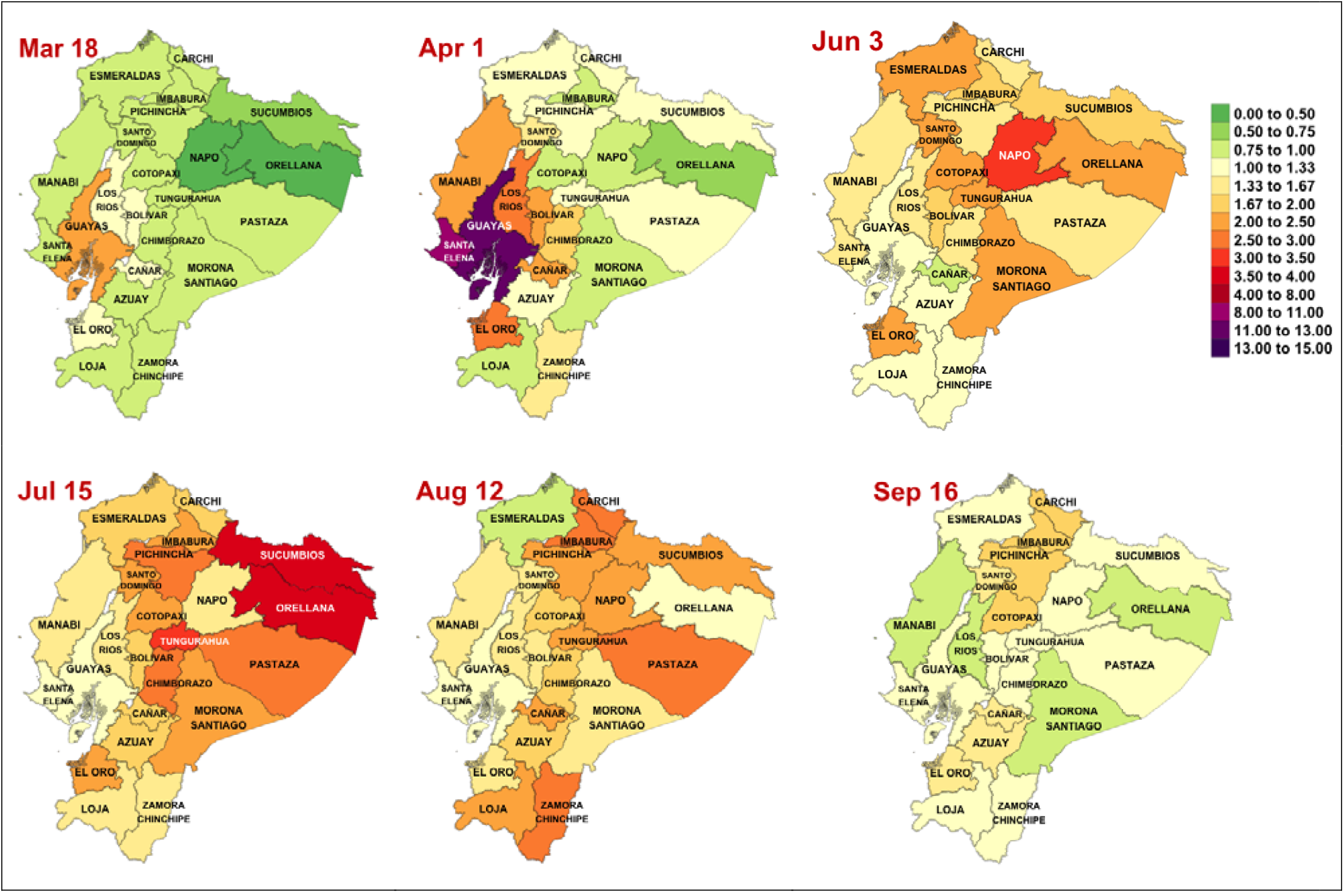
Geographical and temporal evolution of weekly excess death factor for Ecuador

It is easier to visualize the dynamic of the EDFs by displaying as a heatmap (Figure 4) the time series of EDF. The provinces are ordered by the date of the first time a province recorded a doubling in the number of deaths, i.e., has an EDF of two. The box indicates the period of strict lockdown. The plot clearly shows that COVID-19 did not hit uniformly Ecuador, and that the epidemic exhibits both a temporal and geographic pattern. Interestingly, each province shows a peak period of excess death of about a month or two, during which there are over 2 times more deaths than expected. After that peak period, the excess death ratio declines, but typically remains larger than one. The two vertical black lines indicate the start and end of the strict lockdown, and the red line the start date of mandatory mask use [17].

There is a delay of several weeks between the time of infection and the time of death [37]. As a result, for some provinces such as Guayas and Santa Elena, the lockdown occurred too late in the course of their outbreak to impact the time and magnitude of the peak of excess deaths. Other provinces, such as Santo Domingo de Los Tsachilas and Pichincha benefitted from the strict lockdown as they achieve their peak well after the strict lockdown was lifted. Finally, some provinces, such as Manabí, achieved their peak in the second half of the strict lockdown and possibly benefitted from it.

### 4.2 Geographical distribution of mobility

Figure 6 shows a similar heatmap for the median mobility score. The two black lines indicate the date of the beginning and end of the strict lockdown. The red line is the date of the mandatory mask order. The plot shows that all the provinces decreased their mobility during the lockdown period and exhibit some increased mobility thereafter. After the lockdown, the population in some of the provinces, such as Guayas, Pichincha, and Manabí, maintained a lower mobility than before the lockdown, while the population in other provinces, such as Bolivar and Napo, the mobility returned to, or even exceeded, the level prior to the lockdown.

**Figure 5.**
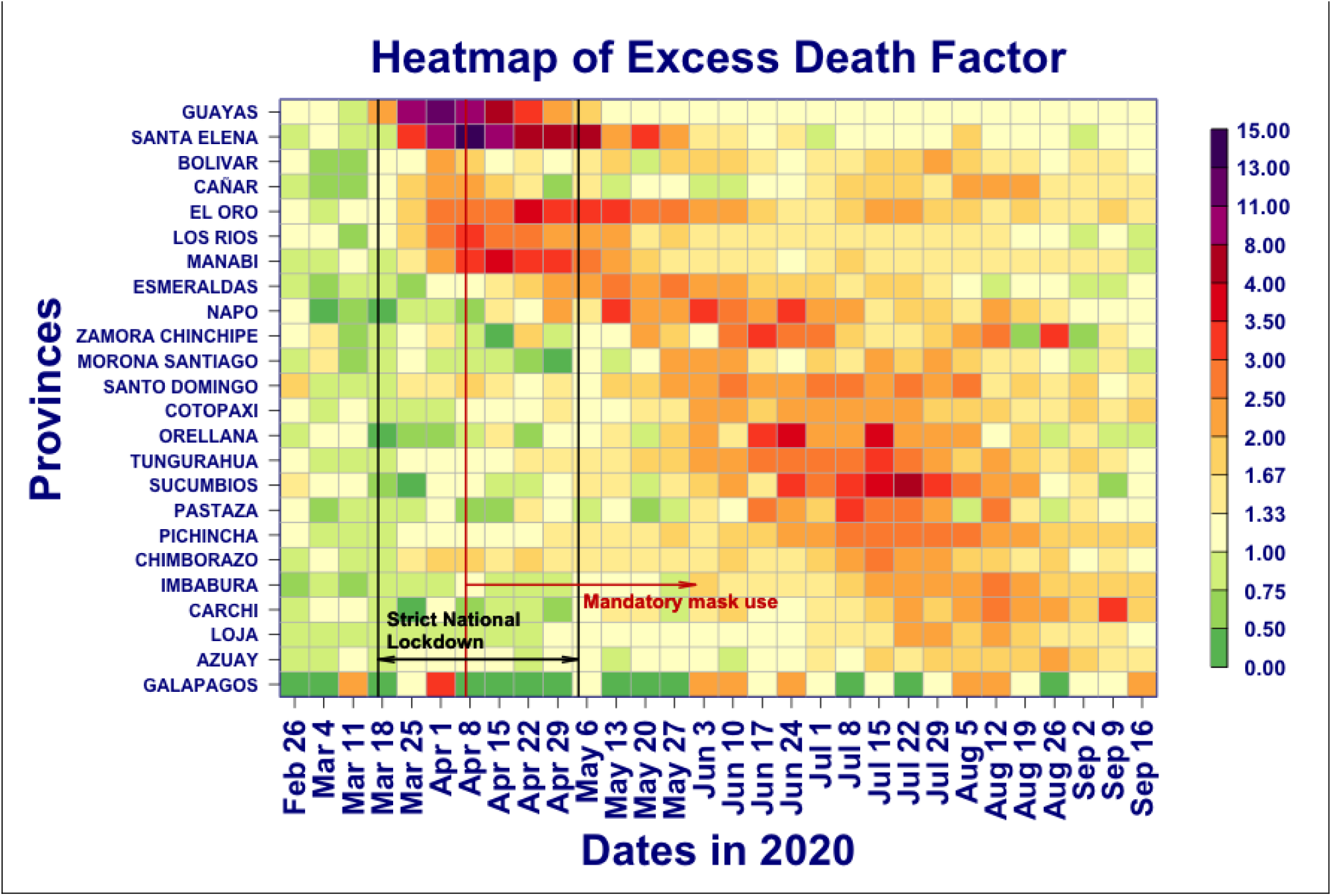
Heatmap of excess death factor by province with national restrictions

**Figure 6.**
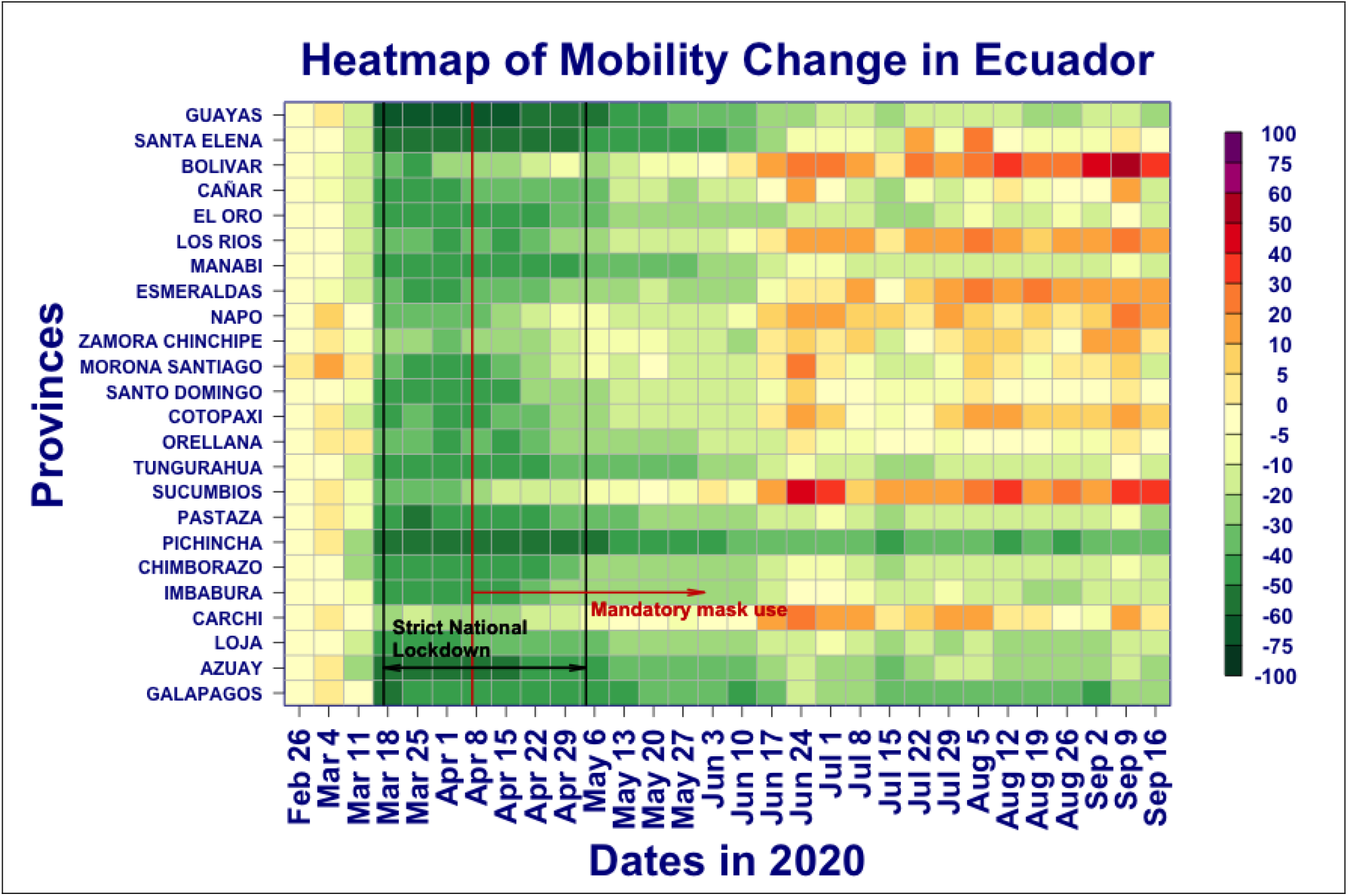
Heatmap of mobility change by province with national restrictions

### 4.3 Relating excess deaths to mobility

Figure 7 display simultaneously the EDF and the median mobility for Guayas, Manabí, Pichincha and Santo Domingo. These four states were chosen because they are archetypes of when they reached their peak of EDF with respect to lockdown period. For Guayas, the lockdown started too late to impact the EDF peak, for Manabí, the lockdown had likely a partial effect on the timing and magnitude of the peak, and both Pichincha and Santo Domingo are examples of peaks occurring well after the lockdown ended. The difference between these two provinces is the pattern of population mobility. In Pichincha, the population maintained a lower mobility than prior to the lockdown, whereas in Santo Domingo, the population mobility returned to pre-lockdown levels. Figure 7, which shows simultaneously the time series of EDFs (in yellow), fraction of deaths attributed to COVID (red) and median mobility score (green) reveals that there is no general pattern linking mobility score to excess death that holds across all provinces. As a result, we performed a separate statistical analysis for each of the 24 provinces, and then compare the results in terms of the stage of the outbreak each province was when interventions were implemented.

**Figure 7.**
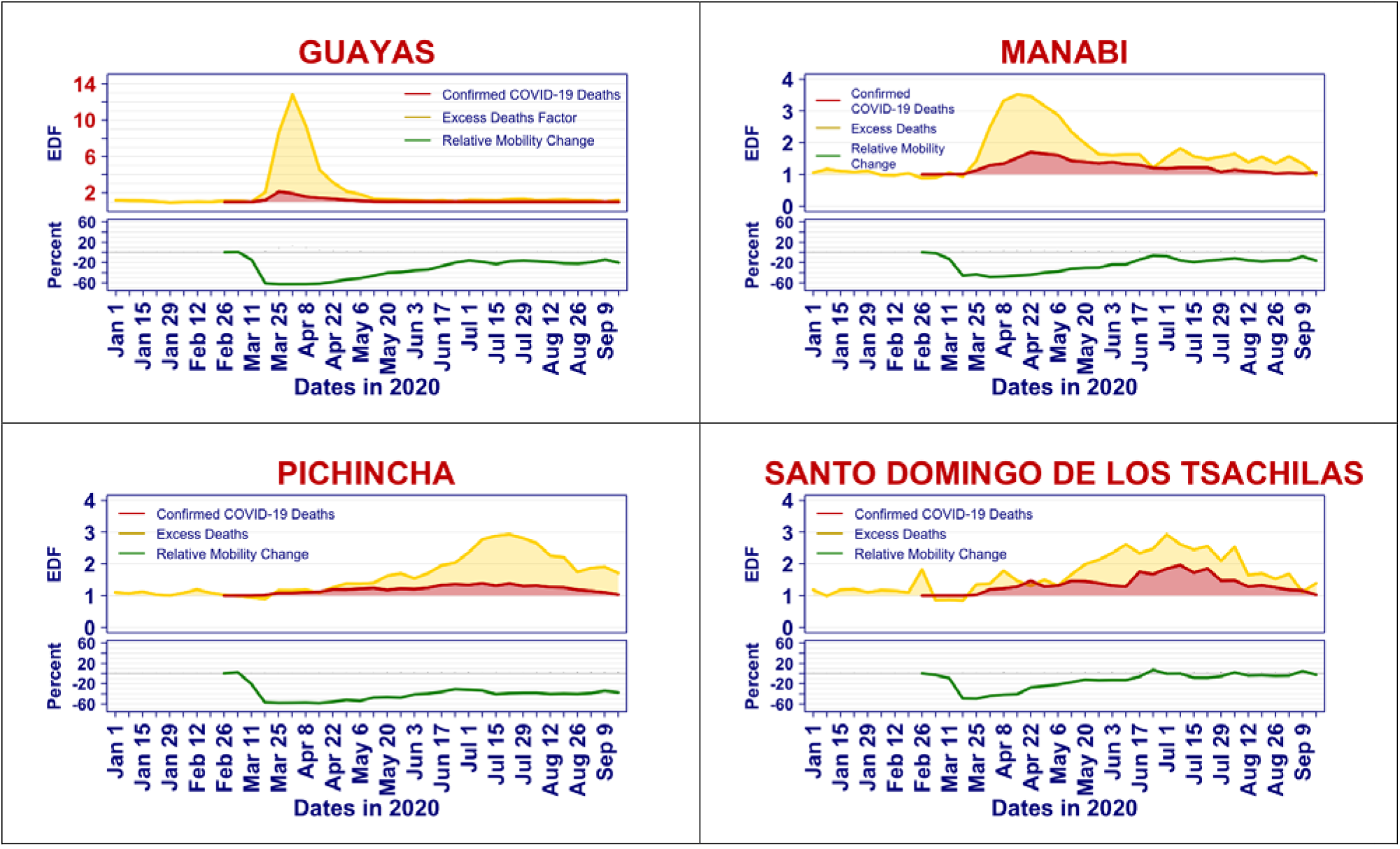
Time series of excess death factor, confirmed COVID-10 deaths, and median mobility

#### 4.3.1 Cross-correlation analysis

Figure 8 displays the cross-correlation between the time series of EDFs and mobility statistics for selected provinces: Guayas, Manabí, Pichincha, and Santo Domingo. The x-axis represents the lag, the difference in weeks, between the mobility statistic and the EDF. A lag of zero (right most point) means the correlation is taken between EDF and the mobility statistic in the same week, whereas a lag of −2 corresponds to a correlation between EDF and the mobility statistic for two weeks prior. The colors indicate positive (green) and negative (red) correlations.

**Figure 8.**
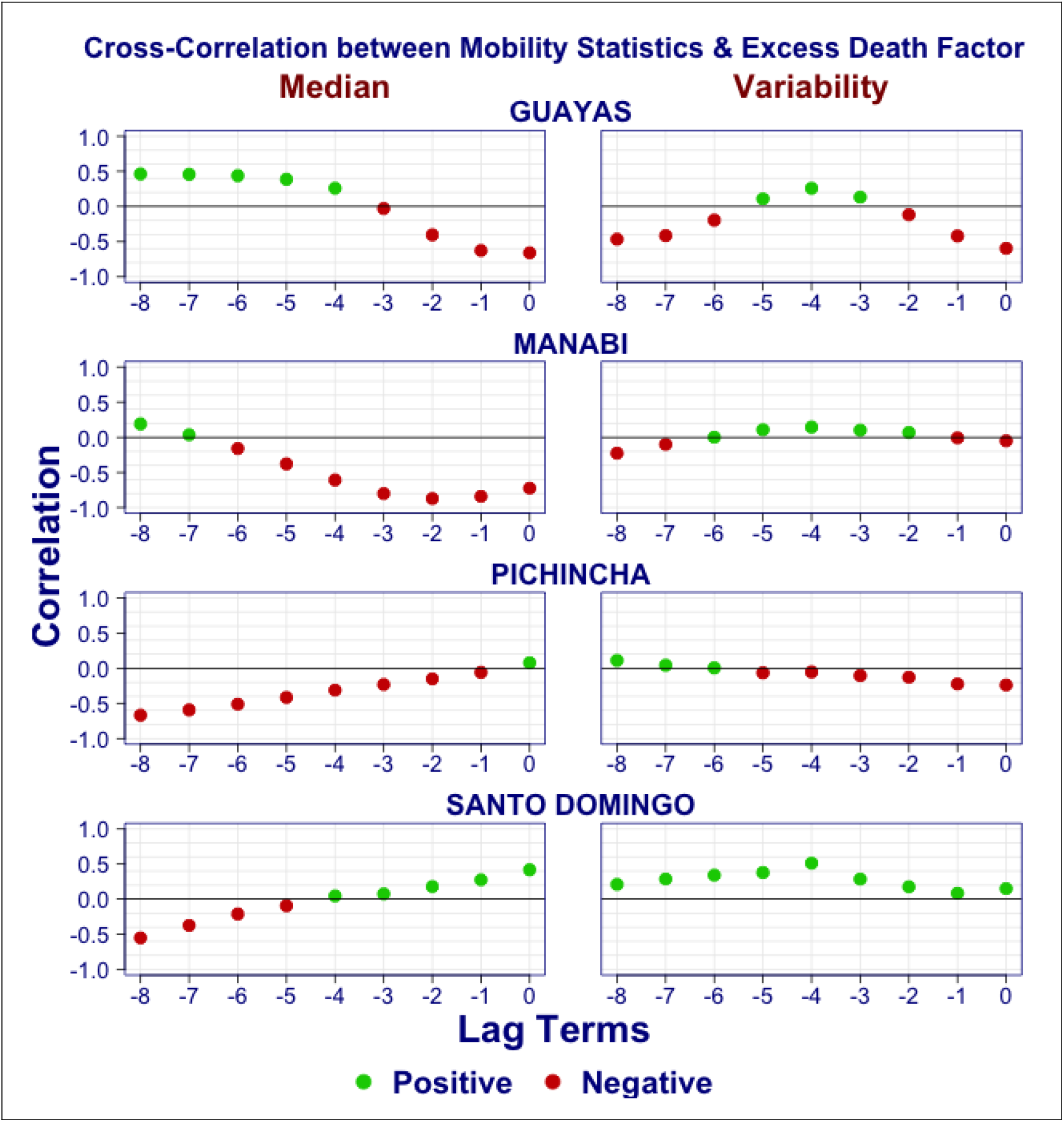
Cross-correlation between time series of excess death factors and mobility statistics

In two of these provinces, Guayas and Manabí, the median mobility scores from the past three weeks are negatively correlated with the current EDF. Looking at Figure 8, we see why: the mobility (green curve) decreases when the EDF (yellow line) is rising. This is indicative of what we expect when a shutdown is instituted because of rising cases.

In contrast, the correlation between EDF and median mobility in the past four weeks is positive in Santo Domingo. Again, **Error! Reference source not found**. tells the story: observe that in Santo Domingo, after the strict lockdown was lifted, the median mobility slowly returned to its pre-lockdown level. At the same time, EDF increased. Again, this is the pattern one expects if one lifts mobility restrictions prior to having fully contained the outbreak.

Finally, in Pichincha, the cross correlation is capturing a situation that lies between the previous cases. After the lockdown ended, the mobility slowly increased, allowing the outbreak to take hold. As it became evident that Pichincha was in the grip of an outbreak, mobility reduced slightly.

Looking at the plot of the cross-correlation for the median in the Appendix, we see the described pattern repeat for all provinces, dividing them into provinces whose outbreak was already in progress at the time of the lockdown, and those whose outbreak emerged after the lockdown.

The interpretation for the variability statistics is harder. Looking at cross correlation plot for the variability for all the provinces (see Appendix), we note that for many provinces, the cross correlation is positive. This makes intuitive sense: larger variability implies that some (but not all) individuals have mobility. For a very infectious virus like SARS-COV-2 [37], a small fraction of individuals who move about and can transmit the disease suffices.

#### 4.3.2 Regression analysis

The regression analysis improves upon the cross-correlation in that it allows to control for age group and sex, considers the joint behavior of a set of explanatory variables, and allows model selection. The latter helps identify which explanatory variables are statistically significant.

We used a log-linear Poisson regression to predict the weekly number of deaths divided into age group and sex categories. By including the logarithm of the predicted death from the baseline model fitted on historical death data as a fixed offset, we can interpret the regression as a model for the excess death ratio. Our model used age group and sex and their interaction as covariates. The inclusion of the age-sex interaction allows the model to fit different death rates for men and women in the same age group. In addition to the demographic variables, we included the value of the median mobility and IQR from the past two to four weeks (names lag 2, lag 3 and lag 4). We applied stepwise regression using the AIC criteria to select which variables are statistically important.

We did not include mobility statistics from the same week or from the one week before to help with the interpretation. Indeed, the mobility statistics of the current and past weeks are correlated, and our model selection could identify those variables as statistically significant, even though we know from the disease progression, and how mobility impacts the spread of disease, that these variables are not causative.

The table of estimated coefficients are provided in **Error! Reference source not found**. of the appendix. To help interpret, we prefer to present in Table 1 the multiplier for the expected EDF from a 10% decrease. For example, a 10% decrease in the median mobility two weeks prior in the province of Santo Domingo, reduces the expected EDF by a multiplier of 0.905.

**Table 1.** Multiplier for the expected excess death factor from a 10% decrease of mobility statistics Lags

| Jurisdiction | Median |  |  | IQR |  |  | % Var. Explained |
| --- | --- | --- | --- | --- | --- | --- | --- |
|  | Lag 2 | Lag 3 | Lag 4 | Lag 2 | Lag 3 | Lag 4 |  |
| **GUAYAS** | 1.713 | 0.804 | 0.803 | 0.803 | 1.342 | 0.854 | 80% |
| **SANTA ELENA** | 1.448 | 1.174 | 0.825 | 1.078 | 0.799 | 0.786 | 81% |
| **BOLIVAR** | 1.048 |  |  |  |  | 0.875 | 32% |
| **CANAR** | 1.117 |  | 0.914 | 0.859 |  |  | 28% |
| **EL ORO** | 1.182 |  | 1.053 |  |  | 0.972 | 88% |
| **LOS RIOS** | 1.183 |  |  |  |  | 0.885 | 80% |
| **MANABI** | 1.240 | 1.075 | 0.950 | 0.946 | 1.057 |  | 82% |
| **ESMERALDAS** |  |  | 1.192 |  |  | 0.914 | 72% |
| **NAPO** | 0.831 | 1.232 |  | 0.897 | 0.846 | 0.908 | 57% |
| **ZAMORA CHINCHIPE** |  |  |  |  |  | 0.906 | 18% |
| **MORONA SANTIAGO** | 0.885 |  |  |  |  | 0.832 | 38% |
| **SANTO DOMINGO DE LOS TSACHILAS** | 0.905 |  | 1.074 |  |  | 0.751 | 45% |
| **COTOPAXI** | 0.914 |  | 1.069 |  |  | 0.938 | 38% |
| **ORELLANA** | 0.803 |  | 1.281 |  | 0.904 | 0.909 | 35% |
| **TUNGURAHUA** |  | 0.829 | 1.190 | 0.916 |  | 0.765 | 52% |
| **SUCUMBIOS** | 0.887 |  |  |  |  |  | 25% |
| **PASTAZA** |  | 0.786 | 1.257 | 0.845 |  | 0.807 | 39% |
| **PICHINCHA** | 0.945 | 0.834 | 1.289 | 1.106 | 0.932 | 0.856 | 29% |
| **CHIMBORAZO** |  |  |  | 1.092 |  | 0.868 | 29% |
| **IMBABURA** | 0.919 |  |  | 0.899 | 0.890 | 0.905 | 72% |
| **CARCHI** |  | 0.907 | 0.913 |  |  | 0.921 | 46% |
| **LOJA** |  | 0.956 |  |  |  | 0.855 | 63% |
| **AZUAY** | 0.929 |  | 1.043 |  | 0.915 | 0.887 | 25% |
| **GALAPAGOS** | 1.322 | 0.754 |  |  |  |  | 14% |

The empty cells in the table indicate variables that have not been selected. For example, the regression model to predict the EDF in the province of Esmeralda only includes the median mobility score from the four weeks before. The provinces are ordered according to the first date that the EDF was two.

Again we see two groups: the provinces for which multipliers of a 10% mobility reduction in lag 2, or when lag 2 is not significant, in lag 3, is greater than one or is less than one. As we observed in the correlation analysis, the provinces that had early peaks during the strict shutdown all show a factor that is greater than one. Indeed, these provinces had an increase in the weekly EDFs and a decrease in mobility at the same time.

The second group had their peak after the end strict shutdown. As we noted the median mobility was increasing during that time. And as mobility increased so did the EDFs. In contrast, multipliers less than one represent the effect of a 10% decrease in mobility.

We note that for almost all provinces the variability (IQR) four weeks prior that is statistically significant, a 10% decrease in the IQR lead to a reduction in the expected EDF.

Finally, the table presents the measure of variability explained by the mobility statistics defined in the Methods section.

A limitation of our analysis is that it does not account for the dynamic of the outbreak. We plan to present such an analysis in a forthcoming paper.

## 5. Conclusions

This paper presents the Excess Death Factor (EDF) time series for all provinces in Ecuador, and relates them to mobility data derived from cellular phone data that was obtained from GRANDATA and the United Nations Development Program for the period starting on March 1^st^, 2020 and ending on September^23rd^, 2020. The data reveals that the provinces were hit by the pandemic in a clear spatio-temporal pattern, with the peak EDF moving across Ecuador over time in a relatively short six-month period. A statistical analysis reveals that the relationship between human mobility and EDF show two archetypes, one pattern for the provinces whose peak EDF occurred during the strict lockdown, and another pattern for the provinces who reached the peak of their EDF after the conclusion of the strict lockdown. Finally, we demonstrate both the median mobility and the variability of mobility as measured by the IQR are statistically significant predictors for the EDF..

## Data Availability

The data underlying this article cannot be shared publicly due to Ecuador government regulations that the Ecuadorian Ministry of Public Health must approve the research protocol to release reusable COVID-19 datasets. Vital statistics may be available upon request to the Ecuadorian Civil Registry and will be publicly available through the Institute of Statistics and Census according to its statistics calendar. Mobility data used in this paper were provided by the United Nations Development Program (UNDP) and GRANDATA, under the umbrella of UNDP's call for papers "Exploring impact and response to the COVID-19 pandemic in Latin America and the Caribbean using mobility data", to promote policy-oriented research on the COVID-19 pandemic effects in LAC. Findings, interpretations, and conclusions are from the authors and do not necessarily represent UNDP's views. The data may be shared upon reasonable request to the corresponding authors.

