## Supplemental Materials for "Assessing the Impact of Human Mobility to Predict Regional Excess Death in Ecuador"

**Appendix**

**A1. Cross-Correlation between Excess Death Factor and Mobility Statistics**

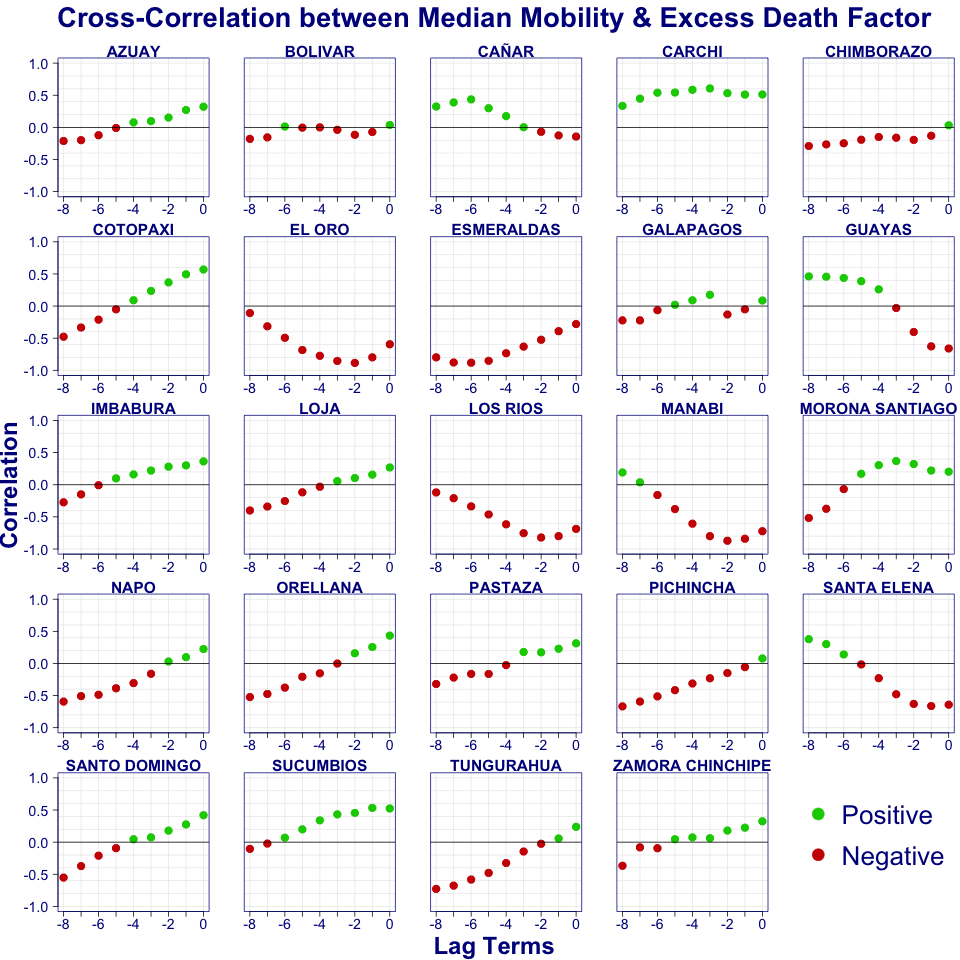

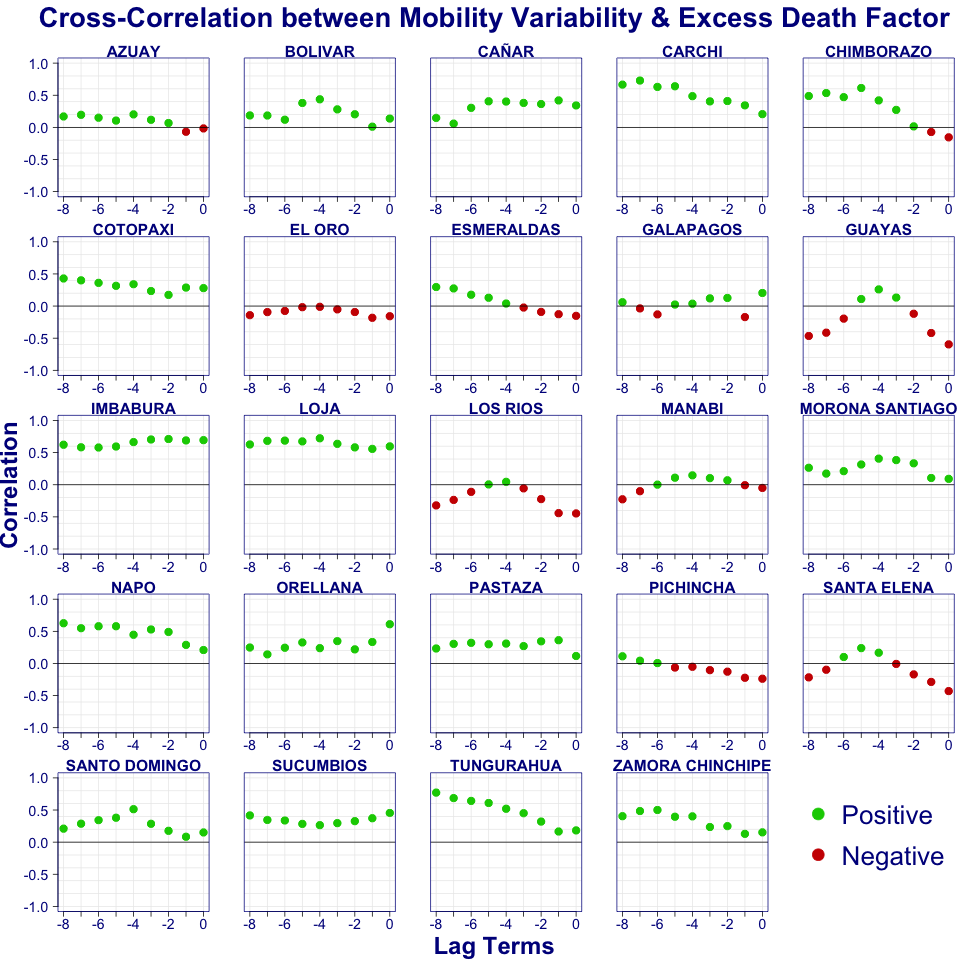

Figure 9 Cross-correlation of excess death factor and median and IQR’s mobility

**A2. Table of coefficient from log-linear regression**

Table 2 displays the estimated coefficient from a Poisson log-linear regression, after model selection. Only the statistically significant coefficients are shown. The provinces are in order of the date of the first time the excess death factor was 2.

Table 2 Regression coefficients of mobility statistics lags

| **Jurisdiction** | **Median** | | | **IQR** | | | **% Var. Explained** |
| --- | --- | --- | --- | --- | --- | --- | --- |
|  | Lag 2 | Lag 3 | Lag 4 | Lag 2 | Lag 3 | Lag 4 |  |
| **GUAYAS** | **-5.38** | 2.18 | 2.19 | 2.19 | -2.94 | 1.58 | 80% |
| **SANTA ELENA** | **-3.70** | -1.60 | 1.92 | -0.75 | 2.25 | 2.41 | 81% |
| **BOLIVAR** | -0.47 |  |  |  |  | **1.34** | 32% |
| **CANAR** | -1.11 |  | 0.90 | **1.52** |  |  | 28% |
| **EL ORO** | **-1.67** |  | -0.52 |  |  | 0.28 | 88% |
| **LOS RIOS** | **-1.68** |  |  |  |  | 1.22 | 80% |
| **MANABI** | **-2.15** | -0.72 | 0.51 | 0.55 | -0.55 |  | 82% |
| **ESMERALDAS** |  |  | **-1.76** |  |  | 0.90 | 72% |
| **NAPO** | 1.85 | -2.09 |  | 1.09 | **1.67** | 0.97 | 57% |
| **ZAMORA CHINCHIPE** |  |  |  |  |  | **0.99** | 18% |
| **MORONA SANTIAGO** | 1.22 |  |  |  |  | **1.84** | 38% |
| **SANTO DOMINGO DE LOS TSACHILAS** | 1.00 |  | -0.71 |  |  | **2.86** | 45% |
| **COTOPAXI** | **0.90** |  | -0.67 |  |  | 0.64 | 38% |
| **ORELLANA** | **2.20** |  | -2.48 |  | 1.01 | 0.95 | 35% |
| **TUNGURAHUA** |  | 1.87 | -1.74 | 0.88 |  | **2.68** | 52% |
| **SUCUMBIOS** | **1.20** |  |  |  |  |  | 25% |
| **PASTAZA** |  | 2.41 | **-2.29** | 1.68 |  | 2.14 | 39% |
| **PICHINCHA** | 0.57 | 1.82 | **-2.54** | -1.01 | 0.70 | 1.55 | 29% |
| **CHIMBORAZO** |  |  |  | -0.88 |  | **1.41** | 29% |
| **IMBABURA** | **0.85** |  |  | 1.07 | 1.17 | 1.00 | 72% |
| **CARCHI** |  | 0.98 | **0.91** |  |  | 0.82 | 46% |
| **LOJA** |  | 0.45 |  |  |  | **1.57** | 63% |
| **AZUAY** | 0.74 |  | -0.42 |  | 0.89 | **1.20** | 25% |
| **GALAPAGOS** | -2.79 | **2.82** |  |  |  |  | 14% |

**A3. Geographical and Temporal distribution of excess death factors**

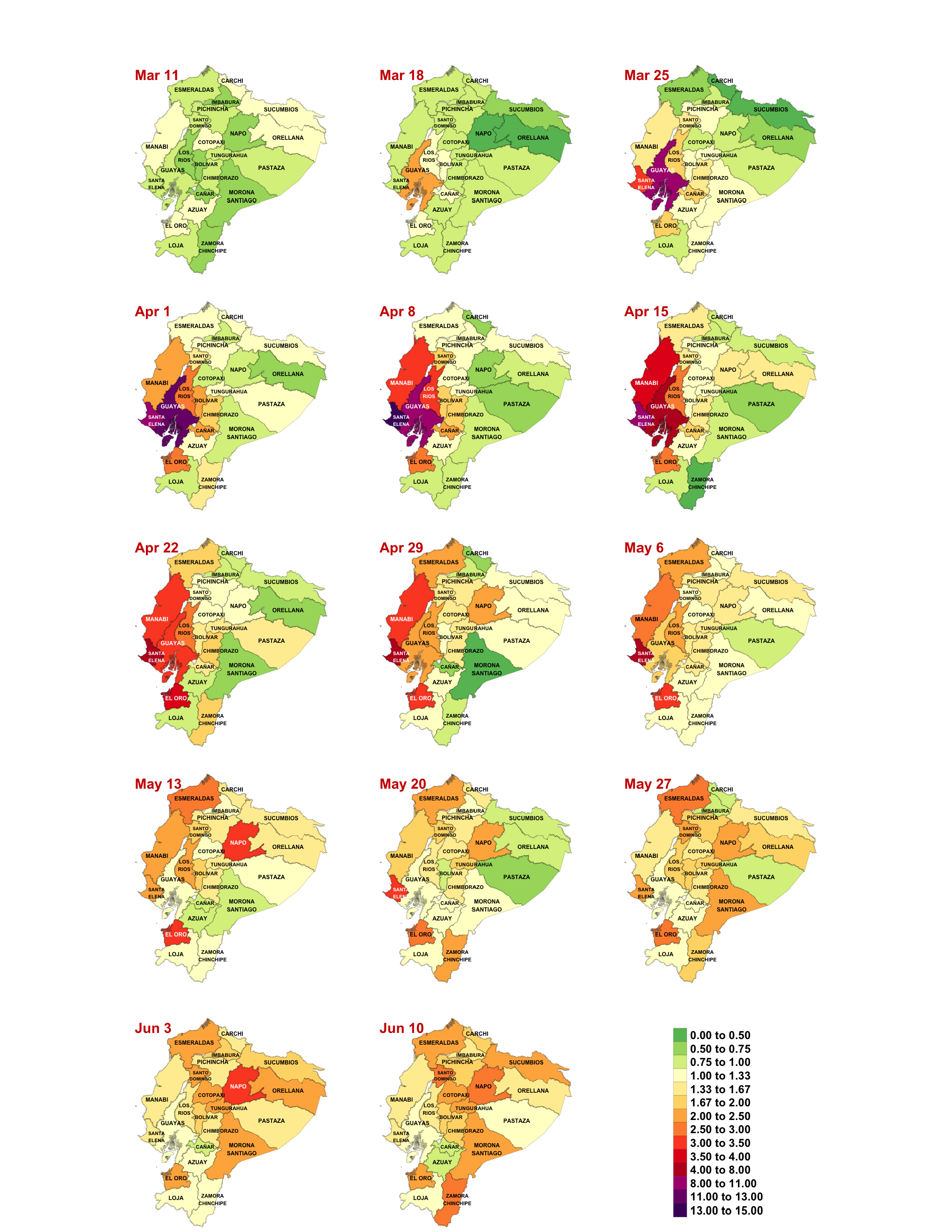

Figure 10 Weekly geographical evolution of excess death factor from March 11 to June16, 2020

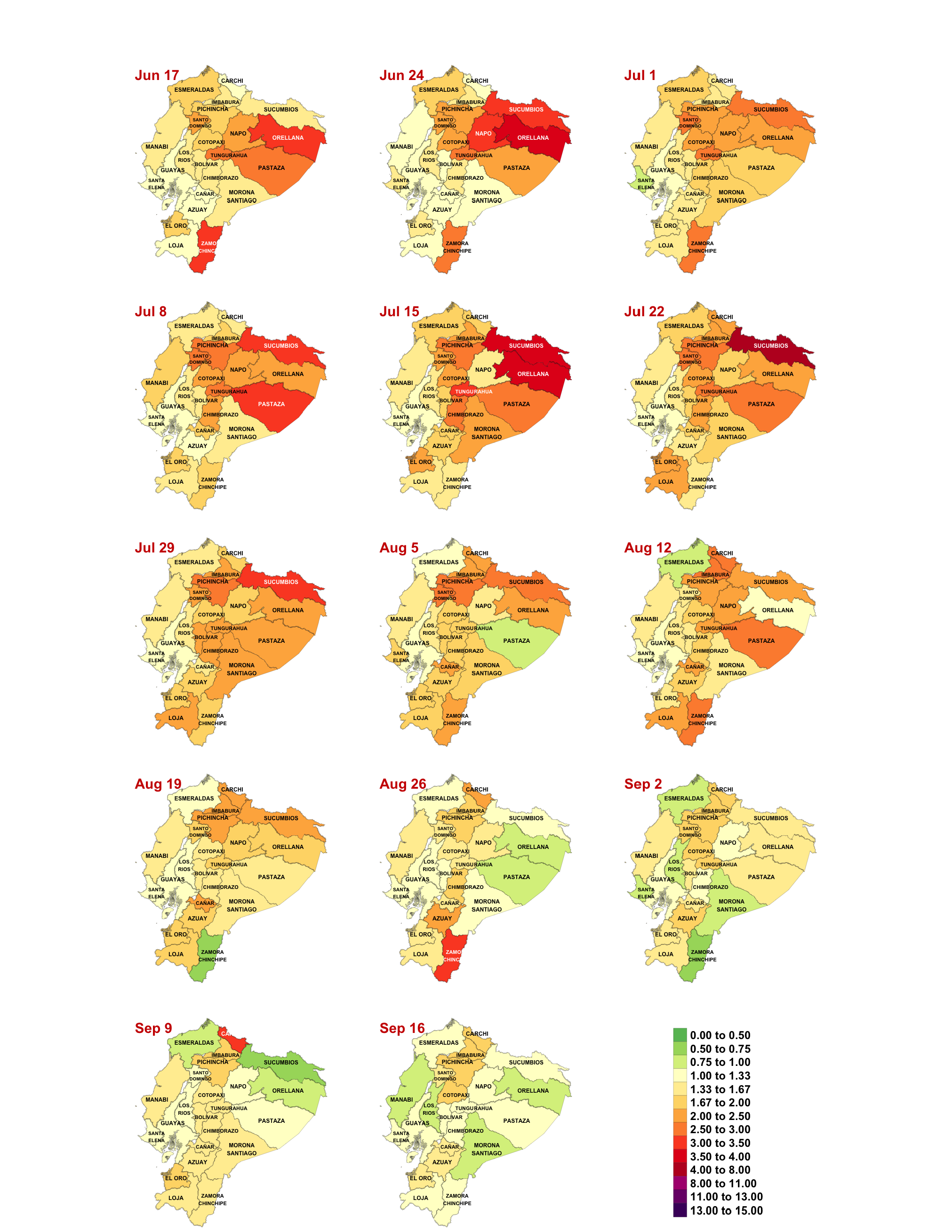

Figure 11 Weekly geographical evolution of excess death factor from June 17 to September 23, 2020
